# Independent Replication and Drug-specificity of an Antidepressant Response Polygenic Risk Score

**DOI:** 10.1101/2022.04.29.22274313

**Authors:** Bochao Lin, Martijn Arns, Bart Rutten, Evian Gordon, Jurjen J. Luykx

## Abstract

We here examine associations of a recently published polygenic risk score of antidepressant response (PRS-AR) with antidepressant treatment outcomes (remission and depression score change) in an independent clinical trial. We not only replicate the PRS-AR for escitalopram, but also find antidepressant interaction effects, suggesting drug-specificity of PRS-AR. We therefore also tested the utility of this PRS-AR to stratify between antidepressants and demonstrate a 14% increase in remission rate (from 43.6% to 49.7%), relative to the randomized remission rate.

## Introduction

Major depressive disorder (MDD) is one of the most common [1], burdensome, and costly psychiatric disorders worldwide. Prescription of antidepressants (AD) is a frequently applied initial step in treating MDD [2], but less than half of individuals achieve remission on their first antidepressant [3]. An evidence-based protocol to individualize antidepressant prescribing to each patient is currently lacking, in part owing to a paucity of established biomarkers [4]. Stratified psychiatry – leveraging biomarkers to stratify between treatments – constitutes a promising step towards precision medicine [5].

Recently, response to antidepressant pharmacotherapy was recognized as a complex, polygenic trait. Several GWASs have been conducted to identify common genetic variants for antidepressant response (AR) [6-10]; yet no genome-wide significant single nucleotide polymorphisms (SNPs) have been identified. Nonetheless, polygenic risk score (PRS) analysis hints that AR is negatively associated with PRS for schizophrenia (SCZ), as well as positively with genetic liability to educational attainment (EA) [7]. The PRS-AR was generated using a meta-GWAS of AR to various antidepressants from multiple cohorts (>80% of patients treated with citalopram/escitalopram). In another set-up, we recently reported that high PRS-SCZ, but not PRS-AR, is associated with better response to ECT in MDD[11], suggesting PRS scores could have utility as stratification markers.

In recent studies we have reported evidence that biomarkers may be drug-class specific (e.g. SSRI vs. SNR) [12], as well as drug-specific [13]. Given the lack of established genetic markers for AR, we conducted a validation study of PRS-AR and examined possible drug-specific associations between PRS-AR, remission and ΔHDRS in an independent, large antidepressants trial not included in the original AR GWAS [7]. We hypothesized that PRS-AR replicates for its associations with a) escitalopram, given that it was the most frequently included drug to generate the PRS-AR; and b) a combined group of several antidepressants. We defined ‘remission’ as primary and change in depression rating (i.e. ΔHDRS) as secondary outcome measures. As a final step we explored the utility of PRS-AR as a stratification biomarker.

## Method

### Participants

Data were collected as part of the International Study to Predict Optimized Treatment for Depression clinical trial (iSPOT-D), a multicenter, randomized, open-lab trial assessing response to three of the most commonly prescribed first-line ADs (for details also see [12] and [14]). The final sample included data for a total of 881 MDD participants recruited from 16 sites in Australia, New Zealand, the USA, and The Netherlands (Supplementary Tables 1 and 2). MDD participants were randomized to escitalopram, sertraline or venlafaxine-extended release. Outcome measures were the 17-item Hamilton Depression Rating Scale change (ΔHDRS) from before to after eight weeks of antidepressant treatment and remission, defined as either an HRSD17 score ≤7 [13].

### Genotyping and quality control

Genotype data for 881 MDD participants was generated on an Affymetrix Direct-to-Consumer (DTC) array containing 497,962 single nucleotide polymorphisms (SNPs). Quality control procedures were performed using PLINK v1.9 [15] using widely accepted, rigorous and published quality control (QC) steps [11] (Supplementary Methods and Supplementary Table 3). In total, 520 European ancestry individuals and 382,258 SNPs passed these QC steps. After merging with the phenotype file, the per-protocol sample consisted of 357 individuals with both genetic and phenotypic information available for subsequent PRS generation and analyses (Supplementary Methods). Additional SNPs were imputed on the TopMed reference panel [16]. Following post-imputation QC (Supplementary Methods), 7,317,868 SNPs remained for PRS calculation.

### Polygenic risk score calculation

We used recent GWASs of AR [7] that employed two levels of outcomes: remission (PRS-AR_Rem:_ remitters as cases and non-remitters as controls) and percentage improvement (PRS-AR_Per_) for PRS calculations using standardized and published methods (Supplementary Methods) [11]. In brief, we constructed PRSs based on effect alleles weighted by effect estimate size, using PRSice2 [17] for 13 GWAS P-value thresholds (Pt): 5×10^−8^, 5×10^−7^, 5×10^−6^, 5×10^−5^, 5×10^−4^, 5×10^−3^, 0.05, 0.1, 0.2, 0.3, 0.4, 0.5 and 1.

### Statistical analyses

Firstly, residuals of PRSs adjusted for age, sex and the first 5 genetic principal components (PCs) were generated using linear regression. Then, using logistic (for remission) and linear (for ΔHDRS) regression models, we tested associations between PRSs and AD outcomes in the whole sample first and then separately for the three AD groups. For remission, baseline-HDRS17 was added as covariate. The model with the optimal p-value threshold (oP_t_) was selected based on the largest R^2^ and smallest p-value from association models (oPt =5× 10^−5^ from the whole sample). Secondly, we explored antidepressant-specific associations between PRS-AR and the outcome measures by adding antidepressants as factor into the full sample model. Third and finally, we stratified individual patients using the median PRS-AR_Rem_ into PRS-high and PRS-low groups, and calculated antidepressant-specific remission rates (N of remitters/N of MDDs) in each AD group to check whether a better clinical outcome was found for the PRS-informed group relative to the treatment-as-usual group. All analyses were conducted in R (version 4.0.5).

## Results

The PRS-AR_Rem_ and PRS-AR_Per_ were generated for 357 MDD patients remaining after QC. In the entire study population, PRS-AR_Rem_ was positively associated with ΔHDRS (oPt =5× 10^−5^, R^2^= 1.77%, β= 0.03, SE= 0.01, P= 0.01; **Supplementary Figure 1A**) and with remission (oPt = 5× 10^−5^, R^2^=1.97, β= 0.23, SE=0.1, P= 0.02, **Figure 1**). Drug-specific analyses yielded a significant, positive association of the PRS-AR_Rem_ with ΔHDRS (oPt =5× 10^−5^, R^2^= 6.53%, β= 0.07, SE= 0.02, P= 0.005; **Supplementary Figure 1B**) and with remission (oPt = 5× 10^−5^, R^2^=8.92, β= 0.52, SE=0.20, P= 0.008, **Figure 1** green line) only for escitalopram (ΔHDRS: **Supplementary Figure 1C&D, Figure 1** blue and red lines). For PRS-AR_Per_ no significant associations were found.

**Figure 1.**
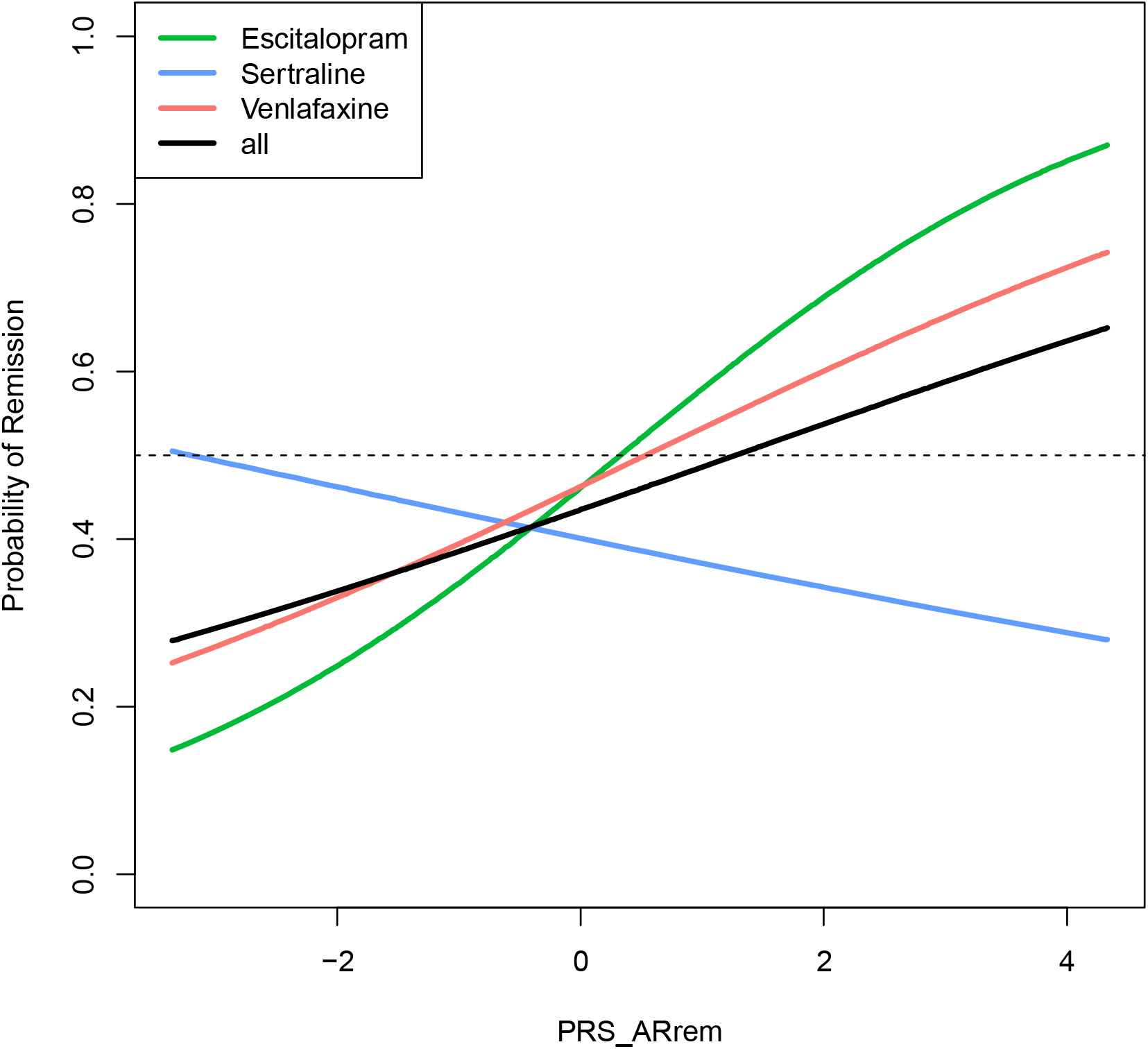
The association between PRS-AR_Rem_ and remission, with antidepressant as a factor in the regression model. Logistic regression of PRS-AR_Rem_ on remission with antidepressant as factor (remission∼ β0 +β1 *PRS + β2*sertraline +β3* venlafaxine+β4*PRS* sertraline +β5* PRS*venlafaxine) was applied. The black curve is the main effect of PRS-AR_Rem_ on remission in the entire study population. The green curve is the PRS-AR_Rem_ effect on probability of remission exp(β1) in the escitalopram group, the blue curve is the PRS-AR_Rem_ effect on probility of remission exp(β1+ β4) in the sertraline group, and the red curve is the PRS-AR_Rem_ effect on probility of remission exp(β1+ β5) in the venlafaxine group. ΔHDRS = the 17-item Hamilton Depression Rating Scale change, PRS-AR_Rem_ = polygenic risk score of remission on antidepressants.

We proceeded to examine the utility of PRS-AR_Rem_ (pt=5× 10^−5^) as a treatment-stratification biomarker between escitalopram, venlafaxine and sertraline. To that end, we added the term ‘antidepressant’ as factor into the full model. Besides the main positive PRS-AR_Rem_ effect on ΔHDRS, there was a significant interaction of PRS-AR_Rem_ with sertraline (β= -0.07, P=0.02; **Supplementary** F**igure 1B** and **Table 1**). Similar results were obtained for remission: besides the main positive PRS-AR_Rem_ association with remission, there was a significant interaction between PRS-AR_Rem_ and sertraline (β= -0.59, P=0.03).

**Table 1.** Regression results of PRS-AR_Rem_ effects with antidepressant interaction effects on ΔHDRS and Remission.

| Outcome | Variables | beta | SE | P |
| --- | --- | --- | --- | --- |
| ΔHDRS | **PRS-AR<sub>Rem</sub>** | **0.067** | **0.023** | **0.003** |
|  | sertraline | -0.024 | 0.036 | 0.502 |
|  | venlafaxine | 0.004 | 0.037 | 0.915 |
|  | **PRS-AR<sub>Rem</sub>\* sertraline** | **-0.074** | **0.033** | **0.024** |
|  | **PRS-AR<sub>Rem</sub>\*venlafaxine** | **-0.034** | **0.032** | **0.293** |
|  | **PRS-AR<sub>Rem</sub>** | **0.490** | **0.190** | **0.010** |
| Remission | **HDRS17-Pre** | **-0.077** | **0.032** | **0.016** |
|  | sertraline | -0.222 | 0.269 | 0.411 |
|  | venlafaxine | -0.014 | 0.276 | 0.959 |
|  | **PRS-AR<sub>Rem</sub>\* sertraline** | **-0.588** | **0.262** | **0.025** |
|  | **PRS-AR<sub>Rem</sub>\* venlafaxine** | **-0.241** | **0.260** | **0.353** |

ΔHDRS = the 17-item Hamilton Depression Rating Scale change
The Model of association between PRS-AR<sub>Rem</sub> and AR outcomes with antidepressant type group as factor (escitalopram as reference group): ΔHDRS/ Remission~ PRS-AR<sub>Rem</sub> + sertraline + venlafaxine + PRS-AR<sub>Rem</sub>\* sertraline+ PRS-AR<sub>Rem</sub>\*venlafaxine. The variables in bold are nominal significant (P<0.05) associated with outcome variables.

Finally, we conducted a simulation study aiming to compare the overall randomized remission rates with PRS-AR_rem_-stratified remission rates. To that end, we performed a median-split of the entire study population into high-PRS (n=174) and low-PRS (n=175) groups. We observed higher remission rates in the high-PRS groups for escitalopram (52.8%) and venlafaxine (52.6%) and in those with low-PRS scores for sertraline (44.1%) than the overall remission rate (43.6%; Table 2).

**Table 2.**
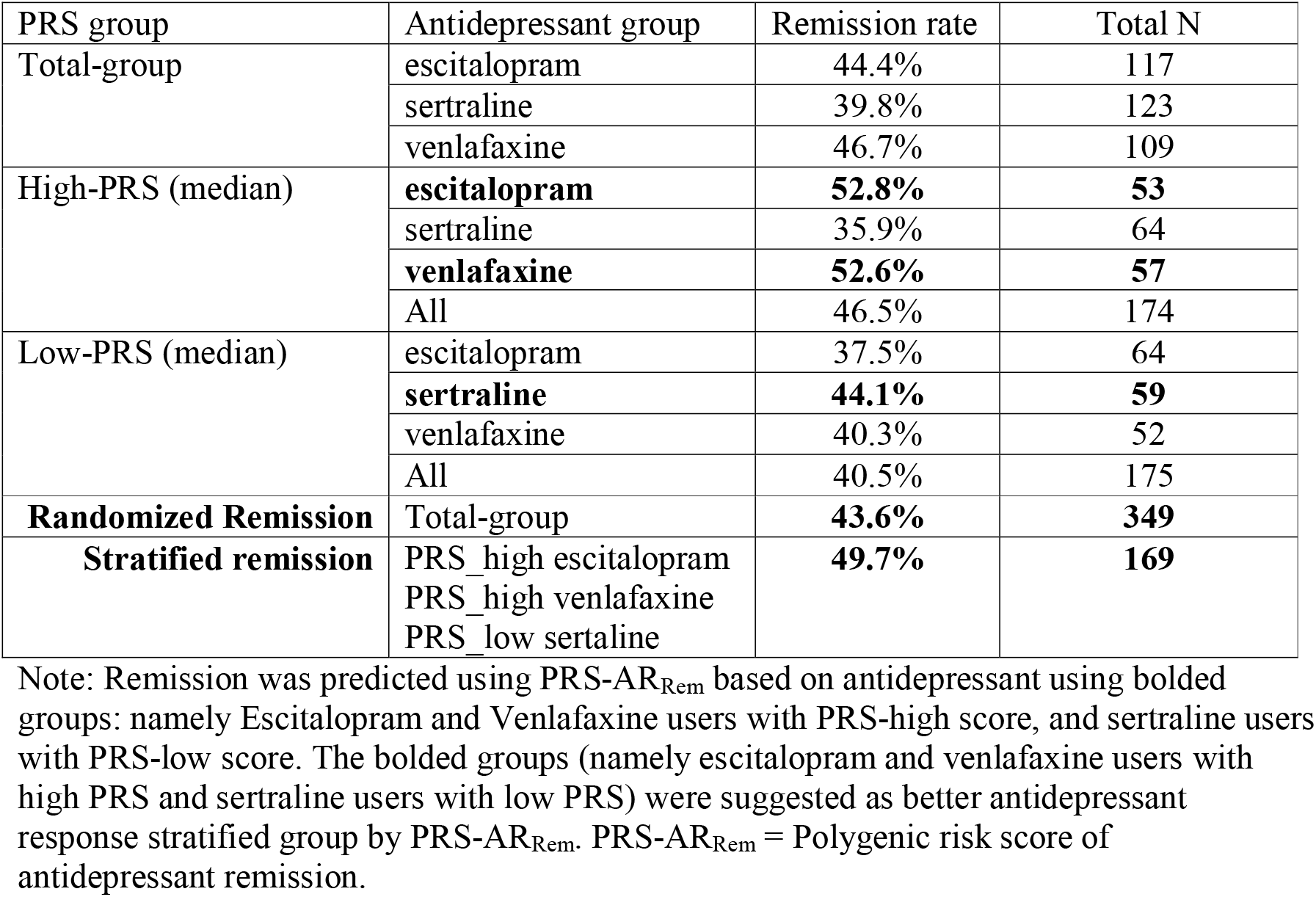
Stratified remission rates based on high vs. low PRS-AR_Rem_.

## Discussion

Here, we demonstrate the first out-of-sample replication for remission and ΔHDRS of the recently reported PRS-AR_Rem_ [7]. When parsed by antidepressant type, this association between remission with PRS-AR_rem_ was only found for escitalopram – which we expected because PRS-AR_Rem_ [7] was generated from studies where >80% of patients had been treated with citalopram/escitalopram, also lending face validity to our replication finding. Follow-up analyses demonstrated a significant interaction when adding drug as a factor to the analyses, with opposing effects for escitalopram and sertraline. While sertraline is also considered a first-line SSRI, sertraline has more pronounced dopamine active transporter (DAT) inhibitory activity, is associated with increased extracellular dopamine in the nucleus accumbens, and striatum, and has anticonvulsant properties that may mediate its antidepressant response [13, 18]. Thus, the current findings strengthen the notion of drug-specific associations between biological variables and outcomes in psychiatry, now also from a polygenic perspective.

We further demonstrated that applying PRS-AR_rem_ may result in a 14% gain in remission rates (from 43.6% to 49.7%), thus hinting at the potential, future utility of employing polygenic risk scores in a clinical context as stratification marker [5]. Future studies should overcome the limitations of this study by additionally examining other PRSs, including for drug-specific effects, and further combining these with other sources of information (e.g. MDD severity, sex, age comorbidities) as well as other biomarkers to try and improve stratification potential and increase remission gains in MDD.

Although our findings hint at better clinical outcomes for the PRS-informed group relative to the randomized treatment allocation, the results await validation in independent cohorts before PRS-informed drug prescribing may be considered in clinical settings.

## Data Availability

All data produced in the present study are available upon reasonable request to the authors.

