## supplementary Material for "Independent Replication and Drug-specificity of an Antidepressant Response Polygenic Risk Score"

### Supplementary Methods

#### Participants

As per the iSPOT-D protocol [1, 2], all participants met criteria for MDD as assessed by the Mini-International Neuropsychiatric Interview in accordance with the DSM-IV [3] and had a score of ≥16 (at least moderate severity) on the Hamilton Depression Rating Scale (HDRS17) [4]. All MDD participants were either antidepressant medication naive or had undergone a wash-out period of at least five half-lives at the start of the study. Exclusion criteria included current or past diagnosis of psychosis, bipolar disorder, posttraumatic stress disorder, obsessive-compulsive disorder, as well as any contraindication to neuroimaging [1, 2]. The study was conducted in accordance with the principles of the Declaration of Helsinki 2008 and written informed consent was obtained from all participants.

**Supplementary Table 1.** Characteristics of iSPOT genotyped participants by recruitment site.

| recruitment centers | Total N | N female (%) | Age (mean±SD) | Education  (mean±SD) |
| --- | --- | --- | --- | --- |
| Brain Dynamics Centre - Australia | 137 | 66 (48.1) | 34.7±13.6 | 14.5±3.0 |
| Flinders University - Australia | 24 | 8 (33.3) | 47.9±10.7 | 12.7±2.5 |
| Swinburne University - Australia | 10 | 5 (50) | 30.4±11.1 | 14.0±2.7 |
| University of Auckland - New Zealand | 20 | 10 (50) | 45.3±10.3 | 14.0±2.5 |
| Brain Resource Centre 60 Hz - US | 347 | 185 (53.3) | 36.9±12.3 | 14.9±3.0 |
| Neurodevelopment Center 60 Hz - US | 11 | 6 (54.5) | 44.0±9.4 | 13.0±1.6 |
| University of Missouri 60 Hz - US | 10 | 6 (60) | 38.8±11.8 | 14.3±1.5 |
| Brain Health South Africa - South Africa | 9 | 8 (88.9) | 33.1±13.4 | 14.1±2.5 |
| Brainclinics Diagnostics BV - Nijmegen, Netherlands | 21 | 12 (57.1) | 47.5±13.0 | 13.5±3.6 |
| Center for Healing the Human Spirit 60 Hz - US | 50 | 28 (56) | 43.0±11.5 | 15.1±2.0 |
| The Alfred - Australia | 10 | 4 (40) | 44.0±12.1 | 14.2±2.7 |
| Shanti Clinical Trials 60 Hz | 102 | 78 (76.5) | 38.7±10.8 | 13.2±2.4 |
| Stanford University 60 Hz - US | 12 | 7 (58.3) | 42.9±15.1 | 14.5±2.4 |
| ADD Treatment Center 60 Hz US | 14 | 8 (57.1) | 46.8±12.6 | 14.9±1.8 |
| Ohio State University 60 Hz - US | 30 | 22 (73.3) | 40.5±14.9 | 15.3±2.4 |
| The Brain Resource Centre 60 Hz - US | 74 | 42 (56.8) | 35.7±11.6 | 15.1±2.9 |
| Total | 881 | 495 (56.2) | 38.2±12.7 | 14.5±2.8 |
| ANOVA test |  | F=11.9; **P=0.0059** | F=3.41; P=0.065 | F=0.26;  P=0.61 |

#### Genotyping and quality control (QC)

Quality control procedures were performed using PLINK v1∙9 according to established methods.[5-7] SNPs and samples with call rates below 95% (n=11) and 98% (n=19), respectively, were removed. Next, strand ambiguous (n=1478) and duplicate SNPs (n=18,610) were removed. A strict SNP QC only for subsequent sample quality control steps was conducted. This involved a minor allele frequency (MAF) threshold >10% and a Hardy-Weinberg equilibrium (HWE) p-value>1e-05, followed by linkage disequilibrium (LD) based SNP pruning (R²<0·2). This resulted in ~54K SNPs to assess heterozygosity (F<3 standard deviation (SD); n=16), homozygosity (F>3SD; n=0), and duplicates (pi-hat>0,8; n=1) and cryptic relativeness (0.125 < pi_hat <0.35; n=40) by pairwise identity by descent (IBD) values. After principal components analysis (PCA) with Hapmap Phase 3 individuals to check ethnicity, samples that deviated more than 10 standard deviations of first 4 PCs from Hapmap 3 European cohorts (n=274) and within our own dataset were removed (n=12). In addition, the first 20 genetic PCs of passed quality controlled samples were generated using the strict SNP QC list in EIGENSTRAT.[8] After removing all failing samples, a regular SNP QC was performed (SNP call rate >98% (n=6,474), HWE p>1e-06 (n=12), MAF>1% (n=61841)). In the end, 520 individuals and 382,258 genotyped SNPs passed these abovementioned QC steps.

The pre-imputation QCed SNPs were then imputed on the TopMed server using 97,256 deeply sequenced human genomes after phasing with Eagle v2.4 [9]. 40,410,413 SNPs with INFO score >0.3 were downloaded from TopMed server. Post-imputation QC involved removing SNPs with a MAF <0.01 and HWE <10^-6^ (n=29,835,638), SNPs that had a discordant MAF (MAF differences >0∙15) compared to the reference panel (n=897,056 SNPs), Rsq info score <0∙8 (n=1,396,158), strand ambiguous AT/CG SNPs (n=952,423) and multi-allelic SNPs (n=9,506). The 22 imputed vcf files per chromosome excluding above-mentioned SNPs were converted into plink best guess format data and then merged for all chromosomes. The merged autosomal hard call plink file contained 520 samples and 7,317,868 SNPs used to calculate PRSs. Details of QC steps are also shown in Supplementary Table 2.

**Supplementary Table 2**. Genotype data quality control (QC) details, which including pre-imputation QC, imputation and post-imputation QC.

| Step | SNP start | SNP  end | Subjects  Start | Subjects end |
| --- | --- | --- | --- | --- |
| Update sex and phenotype information for plink format file | 497,962 | 497,962 | 881 | 881 |
| Remove SNPs with missingness > 0.05 | 497,962 | 497,951 | 881 | 881 |
| Remove samples >2% missing genotypes | 497,951 | 497,951 | 862 | 862 |
| Remove SNPs with insertion or deletion | 497,951 | 470,673 | 862 | 862 |
| Remove SNPs that are strand ambiguous | 470,673 | 452,063 | 862 | 862 |
| *Strict SNP QC: remove SNPs with MAF < 10 %, hardy-Weinberg 0.00001 | 452,063 | 215,833 | 862 | 862 |
| Strict SNP QC: extract common SNPs with hapmap3 (1.12million) | 215,833 | 95,289 | 862 | 862 |
| Strict SNP QC: Prune SNPs on 0.2 max. LD | 95,289 | 55,234 | 862 | 862 |
| Strict SNP QC: exclude long LD regions | 55,234 | 54,306 | 862 | 862 |
| Check heterozygosity  ( +_3SD; n=16) | 54,306 | 54,306 | 862 | 847 |
| Check duplicates (exclude pi_hat > 0.8; 1 pair) | 54,306 | 54,306 | 847 | 846 |
| Check siblings (update family membership .4<pi_hat < 0.6; 11 pairs) | 54,306 | 54,306 | 846 | 846 |
| Check unrelatedness: the inbreeding index F>0.2 (n=0) | 54,306 | 54,306 | 846 | 846 |
| Check unrelatedness (pi_hat <0.35 exclude pi_hat > 0.125) | 54,306 | 54,306 | 846 | 806 |
| ±10 SD ethnicity outliers based on PCA plot with HapMap 3 | 54,306 | 54,306 | 806 | 532 |
| ±3SD ethnicity outliers based on PCA plot with own | 54,306 | 54,306 | 532 | 520 |
| Removed all bad samples using the abovementioned strict QCed SNPs and then do regular SNP QC as mentioned below | | | | |
| Only include autosomal SNPs | 452,063 | 450,585 | 520 | 520 |
| SNP QC – missing genotyped SNPs > 2% (n=6478) | 450,585 | 444,107 | 520 | 520 |
| SNP QC – hardy-Weinberg <1e-6 (n=12) | 444,107 | 444,095 | 520 | 520 |
| SNP QC – removing MAF <1% (n=61841) | 444,095 | 382,254 | 520 | 520 |
| Imputation | 382,254 | 40,410,413 | 520 | 520 |
| Maf <0.01 and HWE<1e-6 | 40,410,413 | 10,575,011 | 520 | 520 |
| Maf difference > 0.15 relative to reference genome | 10,575,011 | 9,677,955 | 520 | 520 |
| Ambigous SNPs | 9,677,955 | 8,281,797 | 520 | 520 |
| INFO>0.8 | 8,281,797 | 7,327,374 | 520 | 520 |
| multi-allelic SNPs | 7,327,374 | 7,317,868 | 520 | 520 |

*Note: These following steps until the line ‘Removed – below’ are the QC steps applied to generate a strict quality control SNPs list, which only used to process bad samples exclusion and to calculate PCs.

#### RPRS calculation

We used recent GWASs Antidepressant response [10] with 2 outcomes (remission: AR_Rem_, and percentage improve: AR_Per_) and antidepressant class response [11] with 4 outcomes (a TRD vs NTRD; b SSRI responders vs non-responders; c SNRI responders vs non-responders; d NDRI responders vs non-responders) for polygenic risk score (PRS) calculations. As a QC step for PRS calculation, the SNPs that overlapped between the summary statistics GWAS (training data-set) [10, 12], 1000 Genomes (reference data-set) [13] and our genotype data (target data-set) were extracted. Then, insertions or deletions, ambiguous SNPs, SNPs with minor allele frequency <0.01 and imputation quality (R2) < 0.8 in both training and target data-sets were excluded. To account for complicated linkage disequilibrium structure of SNPs in the genome, these SNPs were clumped in two rounds with PLINK version 1.90 [5] according to previously established methods: round 1 with the default parameters (physical distance threshold 250 kb and linkage disequilibrium threshold (R2) of 0.5); and round 2 with a physical distance threshold of 5000 kb and linkage disequilibrium threshold (R2) of 0.2. Additionally, we excluded all SNPs in genomic regions with strong or complex linkage disequilibrium structures (e.g. the MHC region on chromosome 6; Supplementary Table 3).

The sample overlap between ISPOT data with MDD and GWAS AR and antidepressant class response samples is unlikely because all samples belong to different cohorts. PRSs were created by taking genetic variants up to varying thresholds of significance from GWAS summary statistics [10, 12] and applying a score from these variants, weighted by the associations in the discovery sample, to predict a trait in an independent target sample.

**Supplementary Table 3**. Characteristics and outcomes in iSPOT genotyped samples.

| Variables | **All (n=349)** | **Female (n=207)** | **Male (n=142)** |
| --- | --- | --- | --- |
| age | 41.39 (13.06) | 40.59 (13.40) | 42.54 (12.51) |
| Years of education | 14.77 (2.68) | 14.87 (2.67) | 14.62 (2.69) |
| HDRS17_Pre | 21.16 (3.60) | 21.23 (3.68) | 21.05 (3.50) |
| HDRS17_8wks | 9.46 (5.92) | 9.52 (5.89) | 9.37 (5.99) |
| HDRS17_pc_dif | -0.55 (0.28) | -0.55 (0.27) | -0.55 (0.29) |
| Remission | 43.5% | 44.0% | 43.0% |
| Response | 61.9% | 62.8% | 60.5% |
| SSRI_SNRI | 31.2% | 32.4% | 29.6% |

Note: HDRS17_Pre: 17-item Hamilton Depression Rating Scale from before antidepressant treatment. HDRS17_8wks: 17-item Hamilton Depression Rating Scale after eight weeks of antidepressant treatment. HDRS17_pc_dif: the precentage of -item Hamilton Depression Rating Scale change(ΔHDRS) from before to after eight weeks of antidepressant treatment, which calculated by (HDRS17_8wks- HDRS17_Pre)/ HDRS17_Pre. SSRI_SNRI: the percentage of SNRI (venlafaxine) in total samples. Remission: samples with either an HRSD17 score ≤7 after eight weeks of antidepressant treatment. Response: samples with >50% reduction in baseline symptoms after eight weeks of antidepressant treatment.

**Supplementary Table 4**. 20 Complex-LD regions and long-range LD regions which were excluded from PRS analysis.

| **Chromosome** | **Base pair position**  (start point to end point) |
| --- | --- |
| 1 | 48000000-52000000 |
| 2 | 86000000-100500000 |
| 2 | 183000000-190000000 |
| 3 | 47500000-50000000 |
| 3 | 83500000-87000000 |
| 5 | 44500000-50500000 |
| 5 | 129000000-132000000 |
| 6 | 25500000-33500000 |
| 6 | 57000000-64000000 |
| 6 | 140000000-142500000 |
| 7 | 55000000-66000000 |
| 8 | 8000000-12000000 |
| 8 | 43000000-50000000 |
| 8 | 112000000-115000000 |
| 8 | 8135000-12000000 |
| 10 | 37000000-43000000 |
| 11 | 87500000-90500000 |
| 12 | 33000000-40000000 |
| 20 | 32000000-34500000 |
| 17 | 40900000-45000000 |

**Supplementary Figure 1.** Associations between PRS-AR_Rem_ and ΔHDRS in the entire study population and in 3 antidepressant groups separately.

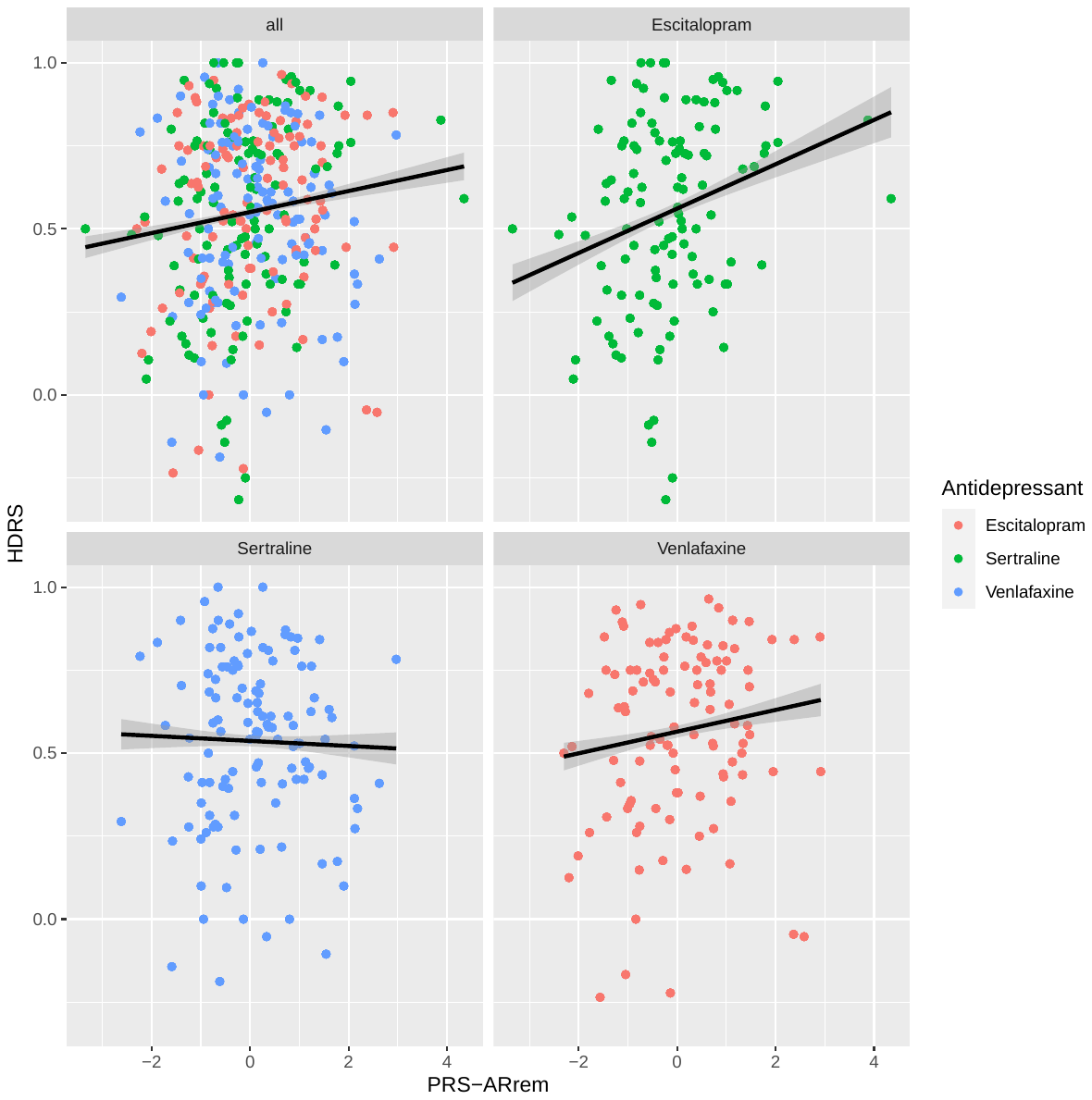

Supplementary Figure 1A. The association between PRS-AR_Rem_ and ΔHDRS in the entire study population (n=357).

Supplementary Figure 1B. The association between PRS-AR_Rem_ and ΔHDRS in escitalopram users (n=117).

Supplementary Figure 1C. The association between PRS-AR_Rem_ and ΔHDRS in sertraline users (n=123).

Supplementary Figure 1D. The association between PRS-AR_Rem_ and ΔHDRS in venlafaxine users (n=109).

The line and grey zones are regression line with 95% confident intervals in linear regression models of PRS-AR_Rem_ on ΔHDRS. Opposing directions of associations can be appreciated for escitalopram and venlafaxine on the one hand and sertraline on the other.

ΔHDRS = the 17-item Hamilton Depression Rating Scale change, PRS-AR_Rem_ = Polygenic risk score of antidepressant remission.
